# Evaluating and Addressing Demographic Disparities in Medical Large Language Models: A Systematic Review

**DOI:** 10.1101/2024.09.09.24313295

**Authors:** Mahmud Omar, Vera Sorin, Reem Agbareia, Donald U. Apakama, Ali Soroush, Ankit Sakhuja, Robert Freeman, Carol R Horowitz, Lynne D. Richardson, Girish N Nadkarni, Eyal Klang

## Abstract

**Background:** Large language models (LLMs) are increasingly evaluated for use in healthcare. However, concerns about their impact on disparities persist. This study reviews current research on demographic biases in LLMs to identify prevalent bias types, assess measurement methods, and evaluate mitigation strategies.

**Methods:** We conducted a systematic review, searching publications from January 2018 to July 2024 across five databases. We included peer-reviewed studies evaluating demographic biases in LLMs, focusing on gender, race, ethnicity, age, and other factors. Study quality was assessed using the Joanna Briggs Institute Critical Appraisal Tools.

**Results:** Our review included 24 studies. Of these, 22 (91.7%) identified biases in LLMs. Gender bias was the most prevalent, reported in 15 of 16 studies (93.7%). Racial or ethnic biases were observed in 10 of 11 studies (90.9%). Only two studies found minimal or no bias in certain contexts. Mitigation strategies mainly included prompt engineering, with varying effectiveness.

However, these findings are tempered by a potential publication bias, as studies with negative results are less frequently published.

**Conclusion:** Biases are observed in LLMs across various medical domains. While bias detection is improving, effective mitigation strategies are still developing. As LLMs increasingly influence critical decisions, addressing these biases and their resultant disparities is essential for ensuring fair AI systems. Future research should focus on a wider range of demographic factors, intersectional analyses, and non-Western cultural contexts.

## Introduction

LLMs are being integrated in multiple sectors, including healthcare (1,2). These models, however, are trained on human-generated text, which often contains biases (3–5). The extent and nature of demographic biases in LLMs remain under-researched. Some studies reveal concerning examples, such as LLMs being less likely to recommend advanced imaging for patients from underrepresented racial groups (6). Similar biases have been observed in legal and other professional domains (7). These biases, influenced by factors like model architecture, training data, and deployment context, can impact critical decisions and have potentially severe consequences (4).

Recent research has shown that commercially available LLMs can perpetuate harmful race-based medical misconceptions (3,5,6). In a study evaluating four LLMs across multiple healthcare scenarios, all models demonstrated instances of promoting debunked racial stereotypes in medicine (8). This can be challenging to detect and measure. Current mitigation strategies include debiasing algorithms, prompt engineering, and diverse training data (9). However, the rapid evolution of these models necessitates ongoing research to ensure future developments promote fairness. This is particularly important given that a recent survey of FDA-approved AI clinical decision support tools found none included a bias evaluation, defined as an analysis to determine whether the tool’s outcomes are fair across different patient groups (10).

We systematically reviewed current research on demographic biases in medical LLMs, aiming to identify prevalent bias types, assess measurement methods, and evaluate mitigation strategies.

## Materials and methods

### Registration and Protocol

We conducted a systematic review following the Preferred Reporting Items for Systematic Reviews and Meta-Analyses (PRISMA) guidelines (11). The protocol was registered with the International Prospective Register of Systematic Reviews (PROSPERO), registration number: CRD42024578467 (12).

### Data Sources and Search Strategy

We searched PubMed, Embase, Web of Science, APA PsycInfo, and Scopus for studies published between January 1, 2018, and July 31, 2024. The search strategy combined terms related to LLMs (e.g., “LLM”, “GPT”, “BERT”) with terms for bias and fairness. We validated our search strings through iterative testing and refinement. We supplemented database searches with manual screening of reference lists from included studies. The full search strategy is available in the **Supplements**. We developed our search strategy following the methods outlined in Chapter 4 of the Cochrane Handbook for Systematic Reviews of Interventions (version 6.4) (13). We used the Rayyan web application for initial screening (14).

### Study Selection

Two reviewers independently screened titles and abstracts using Rayyan software (MO, EK). We obtained full-text articles for all potentially eligible studies. The two reviewers then independently assessed these articles for inclusion. Disagreements were resolved by discussion or arbitration by a third reviewer. The full process is detailed the **Supplements**.

We included peer-reviewed studies that evaluated demographic biases in LLMs applied to medical or healthcare tasks. We defined demographic bias as systematic variation in model outputs based on characteristics such as gender, race, or age (15). We excluded studies of non-LLM models, those focusing solely on model performance without addressing bias, and non-peer-reviewed materials.

### Data Extraction and Quality Assessment

We developed a standardized form for data extraction. One reviewer extracted data, which was verified by a second reviewer. We extracted information on study design, LLM type, types of bias, measurement methods, and key findings. The full process is detailed the **Supplements**.

We assessed study quality using a multi-approach method with the JBI Critical Appraisal Checklist for Diagnostic Test Accuracy Studies and the JBI Critical Appraisal Checklist for Analytical Cross-Sectional Studies. These tools offers a structured framework that can be adapted to assess LLM bias studies, which often share methodological similarities with diagnostic accuracy research. Both fields evaluate outputs against expected standards, examine rates of incorrect classifications, and frequently involve classification tasks. Given the current lack of specific quality assessment tools for LLM bias studies, the JBI checklist provides a flexible approach that can be modified to evaluate crucial aspects such as data selection, bias measurement methods, and control of confounding factors in LLM research.

### Data Synthesis and Analysis

Due to the heterogeneity of included studies, we conducted a narrative synthesis. We categorized studies by type of bias examined, measurement approach, and mitigation strategies proposed. Where possible, we presented quantitative summaries of bias measurements across studies.

## Results

### Search Results and Study Selection

A total of 863 articles were identified through initial screening. After the removal of 257 duplicates and excluding 539 articles through title and abstract screening, 67 articles underwent full-text review. Ultimately, 24 studies met all inclusion criteria (3,6,16–37). A PRISMA flowchart visually represents the screening process in **Figure 1**.

**Figure 1:**
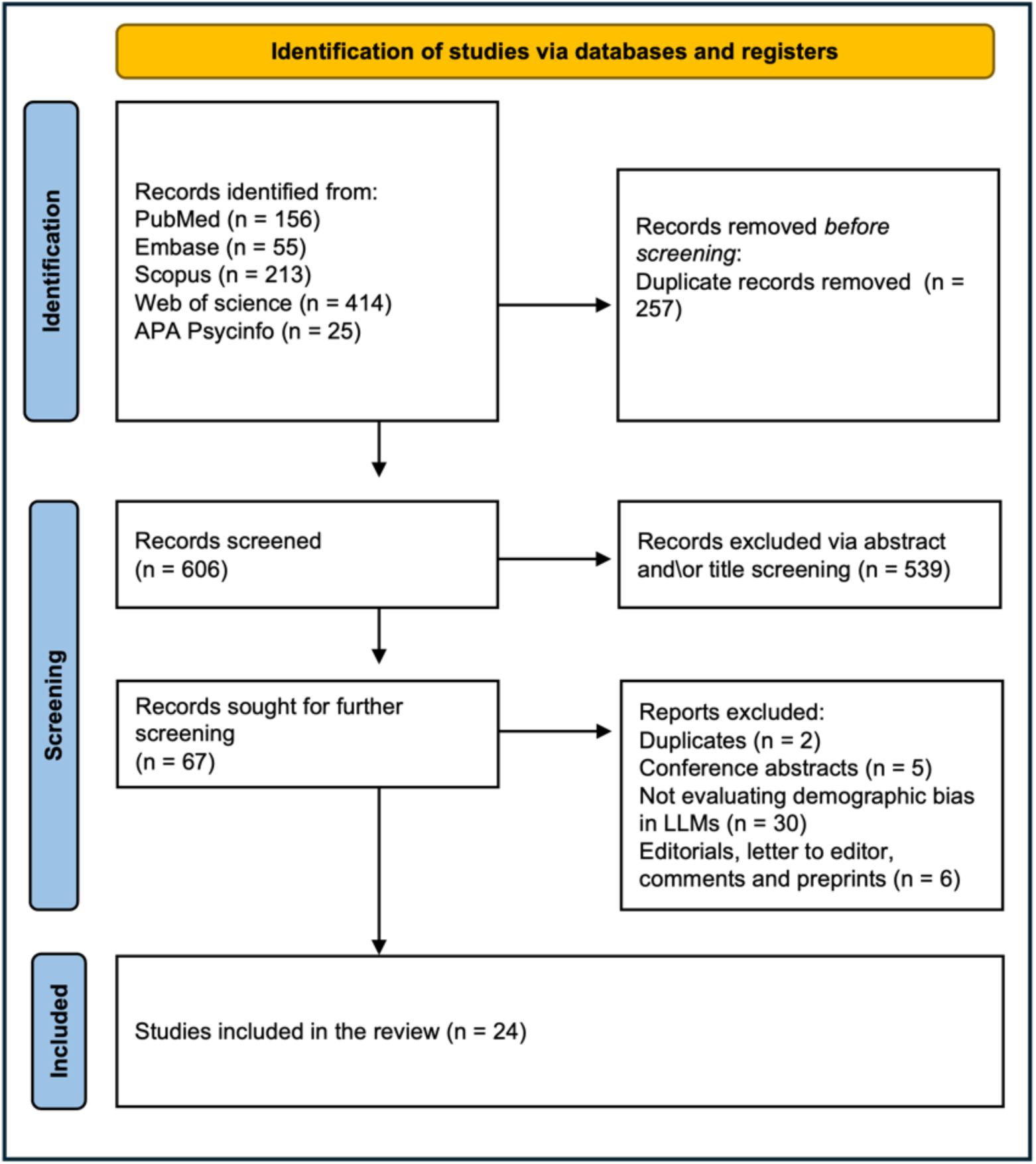
PRISMA flowchart.

### Summary of the included studies

The 24 studies included were published between 2021 and 2024 (3,6,16–37), predominantly from the United States, with contributions from other countries including Germany, the Netherlands, Spain, and Turkey (**Table 1**).

**Table 1:**
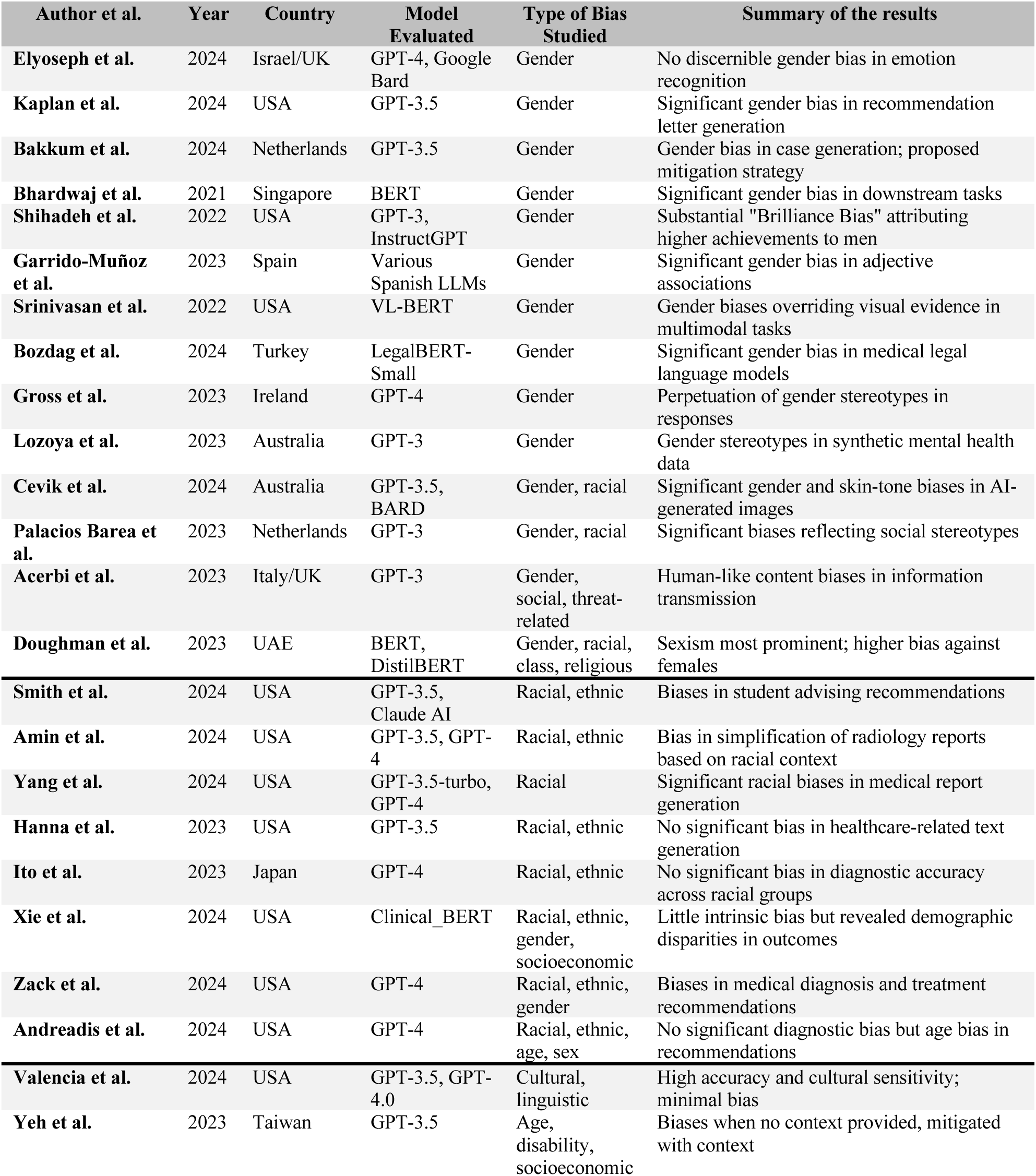
Summary of the characteristics and results of the included studies.

| Author et al. | Year | Country | Model Evaluated | Type of Bias Studied | Summary of the results |
| --- | --- | --- | --- | --- | --- |
| Elyoseph et al. | 2024 | Israel/UK | GPT-4, Google Bard | Gender | No discernible gender bias in emotion recognition |
| Kaplan et al. | 2024 | USA | GPT-3.5 | Gender | Significant gender bias in recommendation letter generation |
| Bakkum et al. | 2024 | Netherlands | GPT-3.5 | Gender | Gender bias in case generation; proposed mitigation strategy |
| Bhardwaj et al. | 2021 | Singapore | BERT | Gender | Significant gender bias in downstream tasks |
| Shihadeh et al. | 2022 | USA | GPT-3, InstructGPT | Gender | Substantial "Brilliance Bias" attributing higher achievements to men |
| Garrido-Muñoz et al. | 2023 | Spain | Various Spanish LLMs | Gender | Significant gender bias in adjective associations |
| Srinivasan et al. | 2022 | USA | VL-BERT | Gender | Gender biases overriding visual evidence in multimodal tasks |
| Bozdag et al. | 2024 | Turkey | LegalBERT-Small | Gender | Significant gender bias in medical legal language models |
| Gross et al. | 2023 | Ireland | GPT-4 | Gender | Perpetuation of gender stereotypes in responses |
| Lozoya et al. | 2023 | Australia | GPT-3 | Gender | Gender stereotypes in synthetic mental health data |
| Cevik et al. | 2024 | Australia | GPT-3.5, BARD | Gender, racial | Significant gender and skin-tone biases in AI-generated images |
| Palacios Barea et al. | 2023 | Netherlands | GPT-3 | Gender, racial | Significant biases reflecting social stereotypes |
| Acerbi et al. | 2023 | Italy/UK | GPT-3 | Gender, social, threat-related | Human-like content biases in information transmission |
| Doughman et al. | 2023 | UAE | BERT, DistilBERT | Gender, racial, class, religious | Sexism most prominent; higher bias against females |
| Smith et al. | 2024 | USA | GPT-3.5, Claude AI | Racial, ethnic | Biases in student advising recommendations |
| Amin et al. | 2024 | USA | GPT-3.5, GPT-4 | Racial, ethnic | Bias in simplification of radiology reports based on racial context |
| Yang et al. | 2024 | USA | GPT-3.5-turbo, GPT-4 | Racial | Significant racial biases in medical report generation |
| Hanna et al. | 2023 | USA | GPT-3.5 | Racial, ethnic | No significant bias in healthcare-related text generation |
| Ito et al. | 2023 | Japan | GPT-4 | Racial, ethnic | No significant bias in diagnostic accuracy across racial groups |
| Xie et al. | 2024 | USA | Clinical_BERT | Racial, ethnic, gender, socioeconomic | Little intrinsic bias but revealed demographic disparities in outcomes |
| Zack et al. | 2024 | USA | GPT-4 | Racial, ethnic, gender | Biases in medical diagnosis and treatment recommendations |
| Andreadis et al. | 2024 | USA | GPT-4 | Racial, ethnic, age, sex | No significant diagnostic bias but age bias in recommendations |
| Valencia et al. | 2024 | USA | GPT-3.5, GPT-4.0 | Cultural, linguistic | High accuracy and cultural sensitivity; minimal bias |
| Yeh et al. | 2023 | Taiwan | GPT-3.5 | Age, disability, socioeconomic | Biases when no context provided, mitigated with context |

*Gender bias* was the most frequently evaluated type (16 studies), followed by *racial and ethnic bias* (11 studies). Other biases examined included *age, disability, socioeconomic status, and sexual orientation*. The studies evaluated various LLMs, including GPT variants (10 studies), BERT variants (7 studies), and other models like ELECTRA and RoBERTa. Methodologies these studies employed for bias detection and measurement varied widely, including prompt-based testing, corpus analysis, task-specific evaluations, and sentiment analysis. Several studies employed statistical techniques such as log-odds ratios, while others used custom metrics or adapted existing frameworks like the Stereotype Content Model (38) (**Table 2**).

**Table 2:**
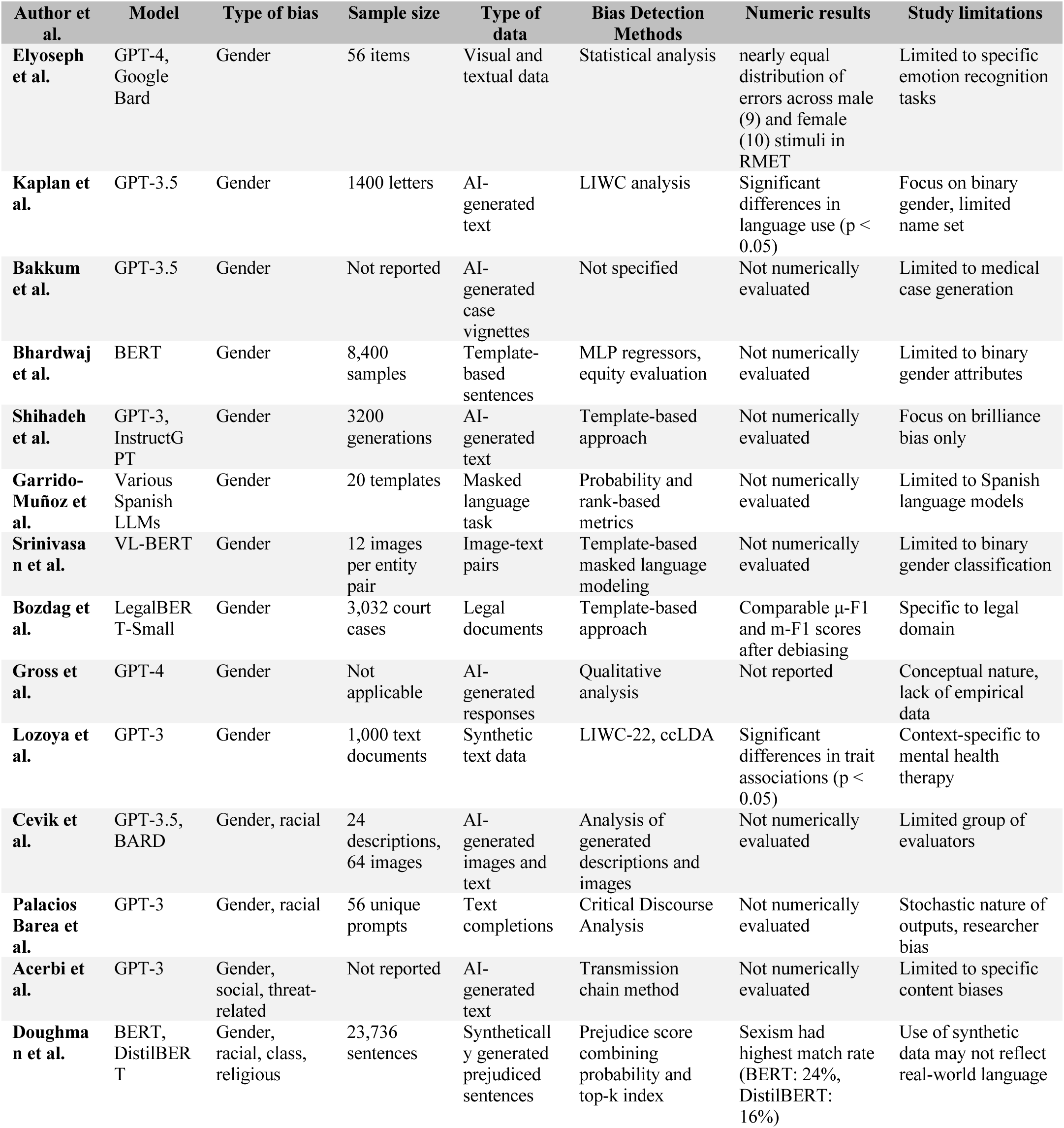
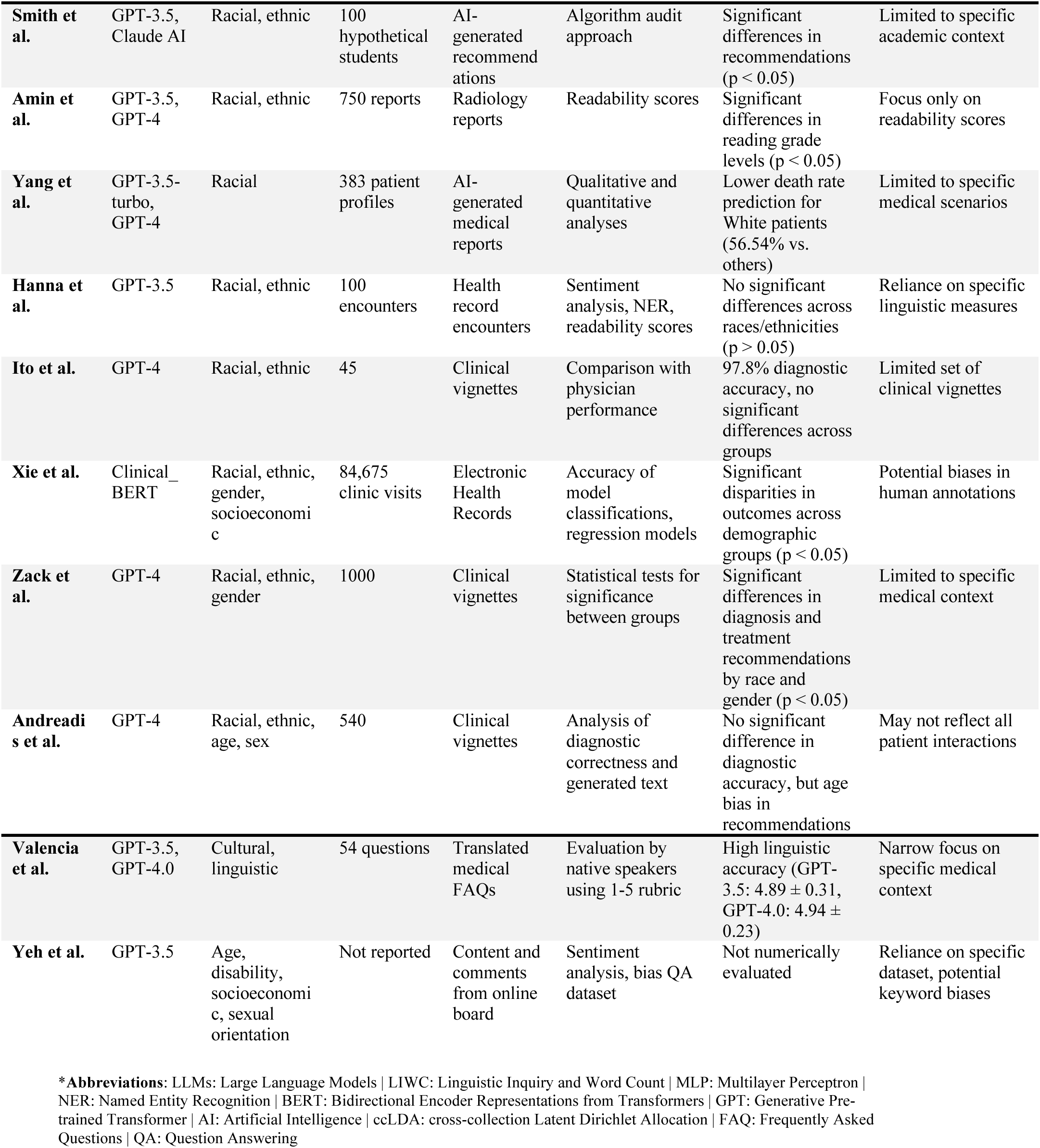
Detailed methodological and quantitative analysis of the included studies.

Out of 24 studies, 22 (91.7%) identified biases in LLMs. Specifically, 15 of 16 studies (93.7%) reported gender disparities, often reflecting traditional gender roles and stereotypes. Additionally, 10 of 11 studies (90.9%) observed racial or ethnic biases, which typically influenced treatment recommendations, language use, or diagnostic accuracy. Pervasive cultural, age, and intersectional disparities were apparent in all evaluated studies (100% of 3, 2, and 3 studies, respectively), while socioeconomic and language biases were noted in 50% of 2 studies each (**Figure 2**).

**Figure 2:**
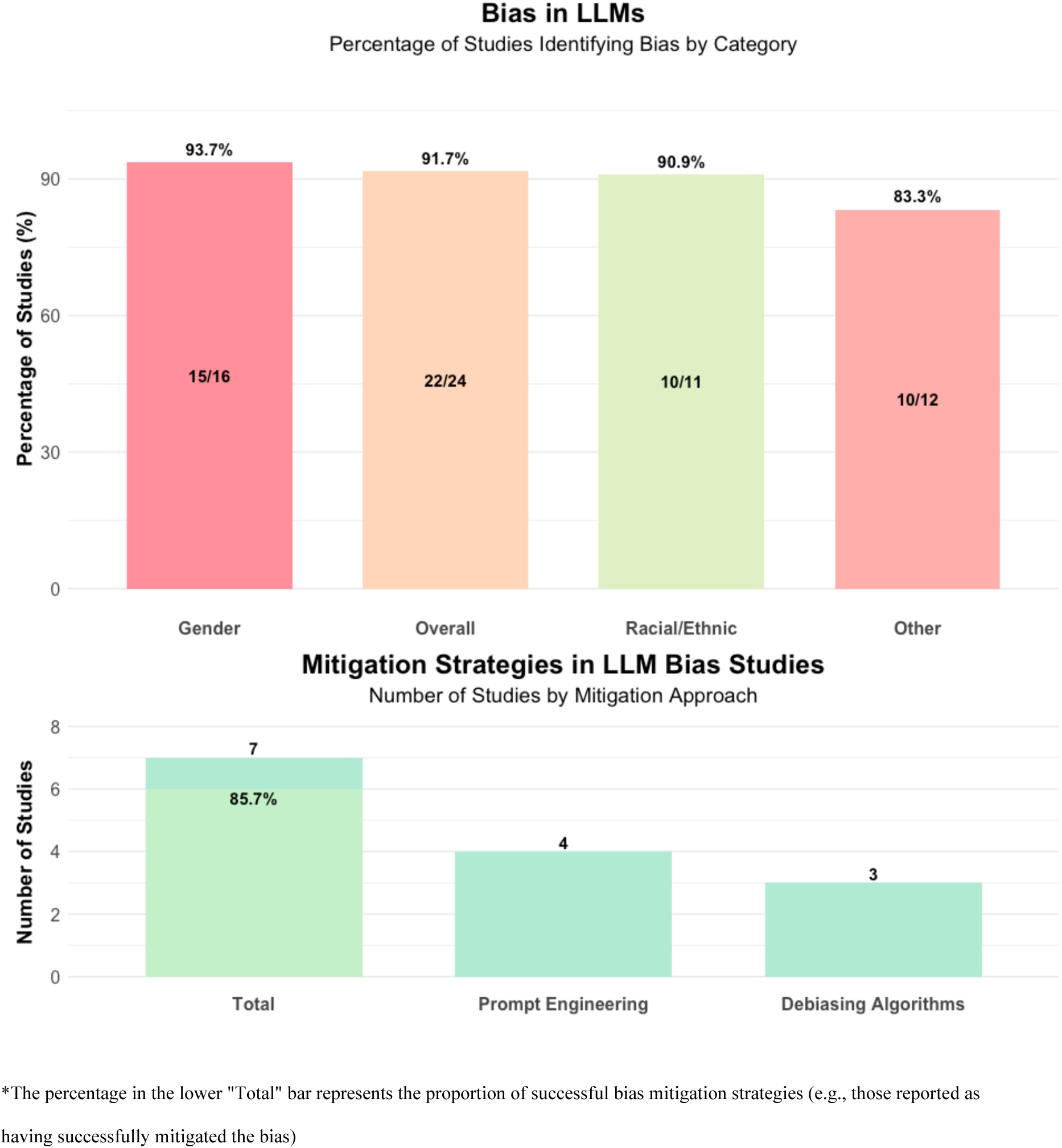
A numeric overall analysis of the detected bias and mitigation strategies.

The studies revealed biases across various LLM tasks in healthcare applications. Newer models like GPT showed demographic bias mainly in text generation tasks, such as creating clinical vignettes and discharge instructions. These models also exhibited bias in prediction tasks, including patient outcome forecasting and diagnostic test recommendations, though to a lesser extent. Older models like BERT displayed bias primarily in classification tasks, with responses differing based on patient race and gender.

Regarding mitigation strategies, 7 studies (29%) implemented explicit methods. Of these, 4 used prompt engineering techniques, and 3 applied debiasing algorithms. Six of the seven studies reported reduced disparities in outcomes after implementing mitigation strategies, showcasing improved fairness in medical applications (**Figure 2**).

### Quality assessment

The quality assessment used two JBI tools: the Critical Appraisal Checklist for Diagnostic Test Accuracy Studies (3 studies) and the Critical Appraisal Checklist for Analytical Cross-Sectional Studies (21 studies) (**Tables S1-2 in the supplements**). Of the 24 studies evaluated, 8 (33.3%) met all applicable criteria. Across all studies, 177 criteria were met (73.8%), 21 were not met (8.8%), 13 were unclear (5.4%), and 29 were not applicable (12.1%). Studies most often met the JBI tools’ criteria related to study design, sample definition, and outcome measurement. Weaknesses included identification and handling of confounding factors, with 7 studies (29.2%) failing to meet or unclear on these criteria. Statistical analysis appropriateness was another concern, with 3 studies (12.5%) not meeting this criterion. The diagnostic accuracy studies generally performed well, meeting most criteria. The cross-sectional studies showed more variability, particularly in addressing confounding factors and statistical analysis.

### Gender bias and mitigation strategies

Gender bias was evaluated in 16 studies across various LLMs and different applications, including GPT variants and BERT variants, with 93.7% confirming its presence. For instance, Kaplan et al., Bhardwaj et al., and Bozdag et al. observed gender bias in text generation tasks (29,32,36). Kaplan et al. found that GPT-3.5 recommendation letters for men included more agentic terms, which describe qualities of assertiveness, independence, and achievement, significantly more than for women who were described using communal language (36). Bhardwaj et al. noted BERT assigned more competence-related traits to male-generated text and more warmth- related traits to female-generated text (29). Bozdag et al. reported gender bias in medical legal contextualized language models affected task performance (32) (**Figure 3**).

**Figure 3:**
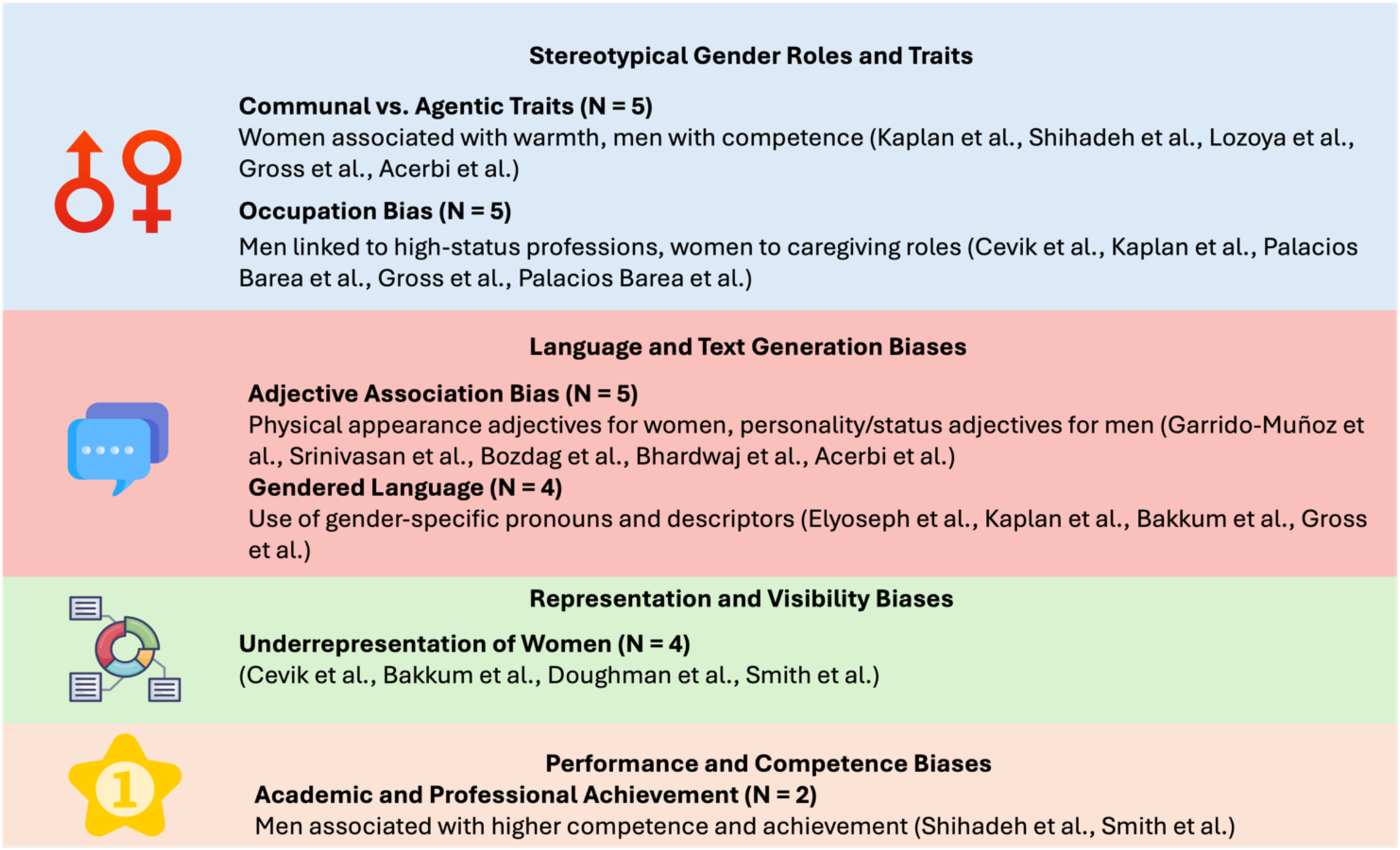
Gender bias manifestations in LLMs.

Bias was also noted in visual tasks. Srinivasan et al. and Gross et al. identified gender stereotypes in visual-linguistic tasks and general responses (3,37). Srinivasan et al. found VL-BERT overrode visual evidence in favor of learned gender biases (37), while Gross et al. reported that GPT reinforced traditional gender roles in its responses (3).

Garrido-Muñoz et al. and Lozoya et al. examined gender bias in non-English contexts (28,31). Garrido-Muñoz et al. found Spanish language models showed strong bias in describing females with body-related adjectives and males with behavior-related adjectives (31). Lozoya et al. observed gender stereotypes in synthetic mental health data generated by GPT-3 (28).

Shihadeh et al., Palacios Barea et al., and Acerbi et al. explored specific aspects of gender bias (20,21,30). Shihadeh et al. found evidence of “Brilliance Bias” in GPT-3 and InstructGPT, attributing higher achievements to men (21). Palacios Barea et al. observed GPT-3 reproduced social stereotypes related to gender (20). Acerbi et al. noted GPT-3 exhibited human-like gender biases in information transmission (30).

There were also some counterexamples as well, as bias mitigation strategies proposed by some authors (**Table 3**). Elyoseph et al. found no discernible gender bias in GPT- 4’s emotion recognition tasks, contrasting with other studies’ findings (22).Valencia et al. demonstrated that prompt engineering could enhance cultural sensitivity in medical translations using GPT-3.5 and GPT-4.0 (16). Similarly, Bakkum et al. proposed a similar prompt engineering method to reduce bias in legal language models while maintaining performance (35).

**Table 3:**
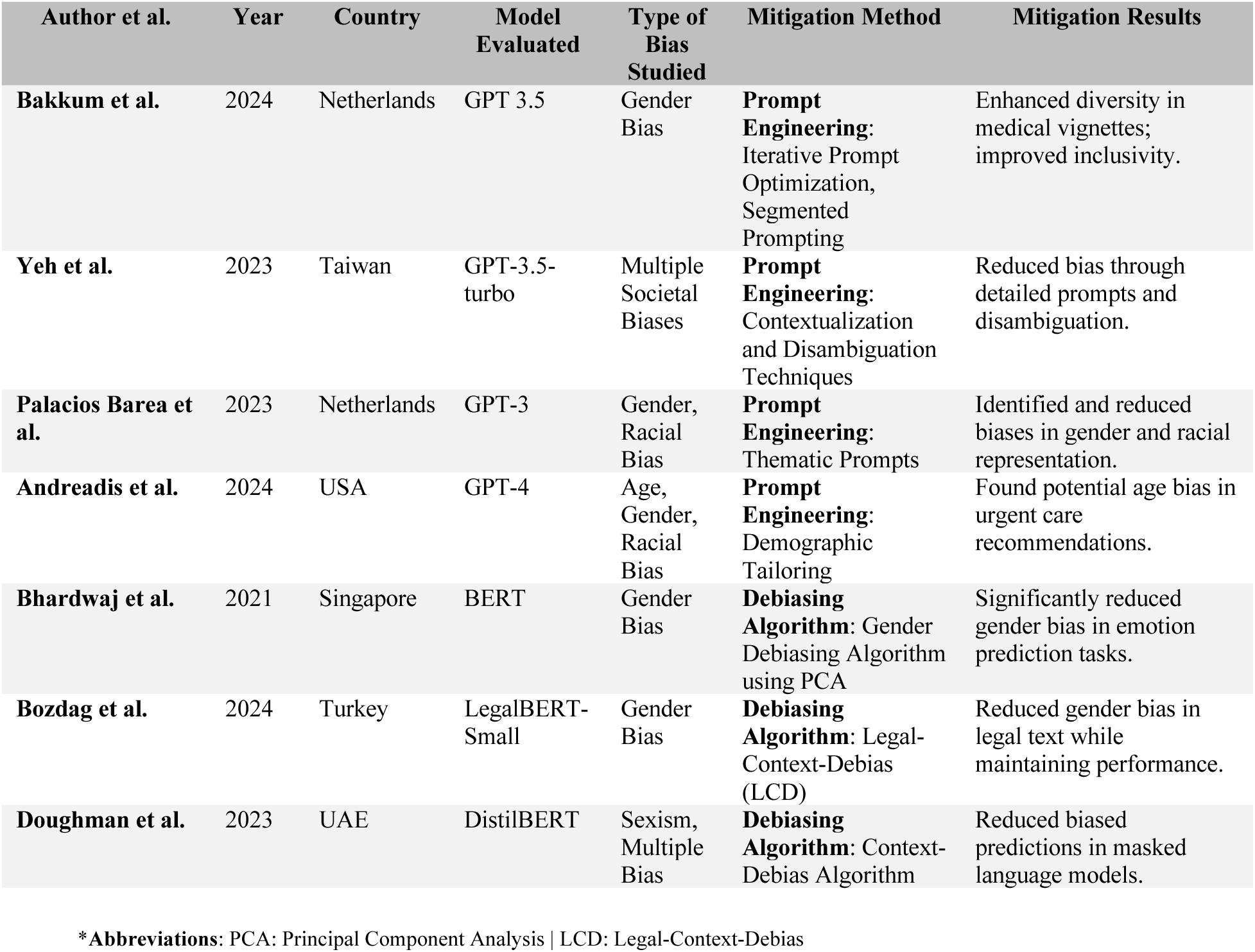
Mitigation strategies reported.

| Author et al. | Year | Country | Model Evaluated | Type of Bias Studied | Mitigation Method | Mitigation Results |
| --- | --- | --- | --- | --- | --- | --- |
| Bakkum et al. | 2024 | Netherlands | GPT 3.5 | Gender Bias | **Prompt Engineering:** Iterative Prompt Optimization, Segmented Prompting | Enhanced diversity in medical vignettes; improved inclusivity. |
| Yeh et al. | 2023 | Taiwan | GPT-3.5-turbo | Multiple Societal Biases | **Prompt Engineering:** Contextualization and Disambiguation Techniques | Reduced bias through detailed prompts and disambiguation. |
| Palacios Barea et al. | 2023 | Netherlands | GPT-3 | Gender, Racial Bias | **Prompt Engineering:** Thematic Prompts | Identified and reduced biases in gender and racial representation. |
| Andreadis et al. | 2024 | USA | GPT-4 | Age, Gender, Racial Bias | **Prompt Engineering:** Demographic Tailoring | Found potential age bias in urgent care recommendations. |
| Bhardwaj et al. | 2021 | Singapore | BERT | Gender Bias | **Debiasing Algorithm:** Gender Debiasing Algorithm using PCA | Significantly reduced gender bias in emotion prediction tasks. |
| Bozdag et al. | 2024 | Turkey | LegalBERT-Small | Gender Bias | **Debiasing Algorithm:** Legal-Context-Debias (LCD) | Reduced gender bias in legal text while maintaining performance. |
| Doughman et al. | 2023 | UAE | DistilBERT | Sexism, Multiple Bias | **Debiasing Algorithm:** Context-Debias Algorithm | Reduced biased predictions in masked language models. |

\*Abbreviations: PCA: Principal Component Analysis | LCD: Legal-Context-Debias

### Racial and ethnic bias

Racial and ethnic biases were examined in 11 studies across several applications. Yang et al. found GPT-3.5-turbo exhibited biases in medical report generation across racial groups, including fabricated patient histories and racially skewed diagnoses (6). Zack et al. reported that GPT-4 showed disparities in recommending advanced imaging, with lower rates of recommendations for patients from underrepresented racial groups compared to those of European descent (18). In a similar manner, Smith et al. found biases in student advising recommendations when examining GPT-3.5 and Claude AI’s responses to lists of names associated with different racial/ethnic groups (27) (**Figure 4**).

**Figure 4:**
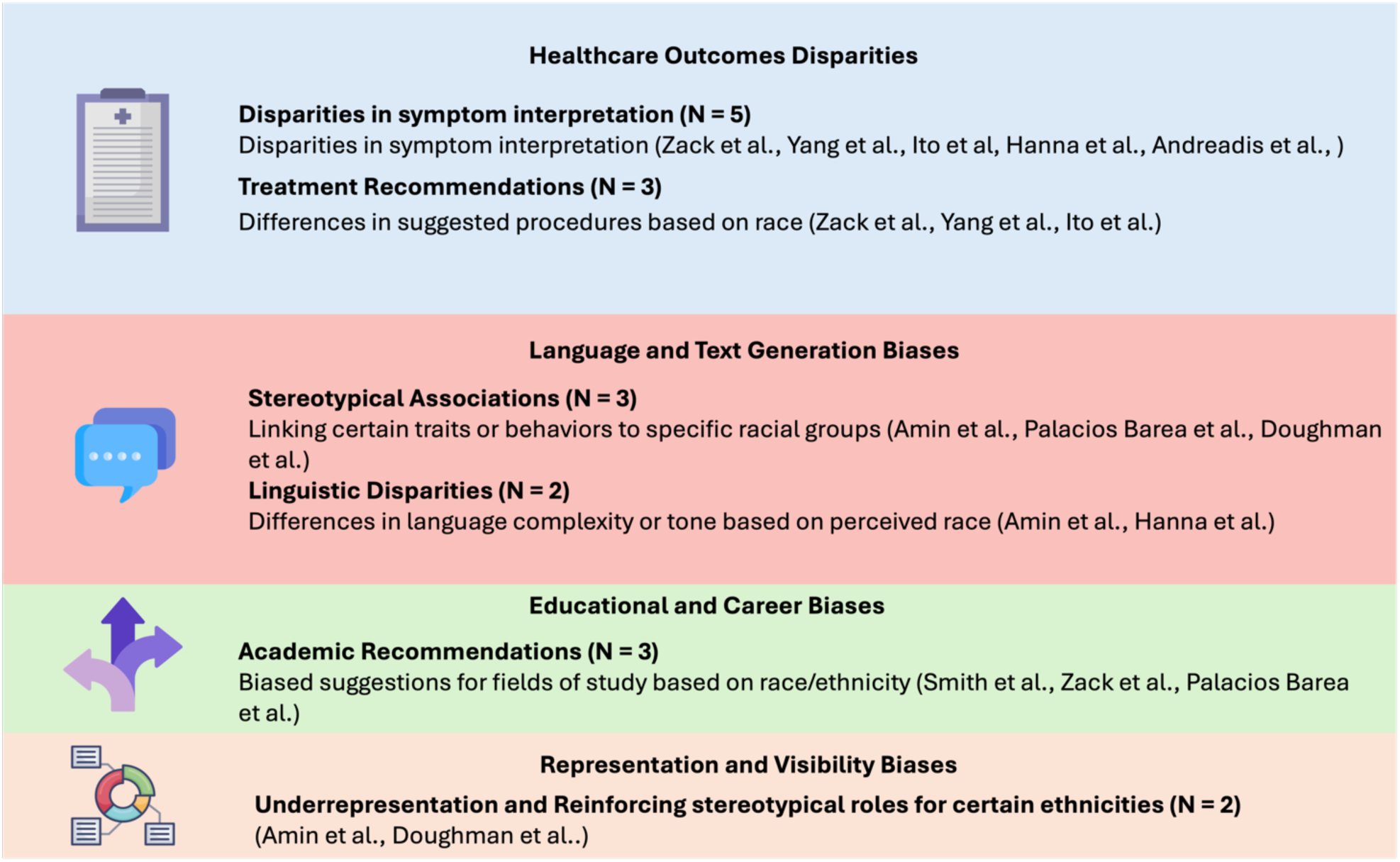
Racial and ethnic biases manifestations in LLMs.

Amin et al. observed bias in GPT’s simplification of radiology reports based on racial context, finding statistically significant differences in reading grade levels between racial contexts for both GPT-3.5 and GPT-4 (25). Conversely, some studies found limited or no evidence of racial bias. Xie et al. observed little intrinsic bias in ClinicalBERT but revealed demographic disparities in outcomes when applied to real-world data (23). Hanna et al. found no significant differences in polarity and subjectivity across races/ethnicities in GPT’s healthcare-related text generation (17). Similarly, Ito et al. reported no significant difference in GPT-4’s diagnostic accuracy across racial and ethnic groups when compared to human physicians (34). Andreadis et al. also reported no significant racial diagnostic bias with GPT-4, although they noted an age-related bias in recommendations (33).

### Other biases

Yeh et al. (2023) conducted a study using GPT-3.5 to examine biases related to age, disability, socioeconomic status, and sexual orientation (24). This research expanded the scope of bias investigation beyond commonly studied gender and racial biases. The study found that GPT-3.5 exhibited biases across these demographic factors when prompts lacked context. However, these biases were mitigated when contextual information was provided (24).

Andreadis et al. observed age-related bias in GPT’s urgent care recommendations, which were presented more frequently to older individuals, even without the proper clinical evidence (33). Xie et al. found socioeconomic disparities in LLM-extracted seizure outcomes, with patients having public insurance and those from lower-income zip codes showing worse outcomes (23). Doughman et al. (2023) conducted a study examining multiple types of bias in BERT and DistilBERT models, including gender, racial, class, and religious biases (26). Their research revealed that sexism was the most prominent form of bias, with a notably higher bias against females. The study found that sexist sentences had the highest match rate, with BERT showing around 24% and DistilBERT showing 16% for sexist content. While the exact definition of sexism used in the study isn’t provided, it likely involved stereotypical or discriminatory representations of women in language. The researchers used synthetically generated prejudiced sentences to evaluate the models, analyzing their predictions on masked tokens (**Table 4** lists some specific examples of different biases from the included studies).

**Table 4:**
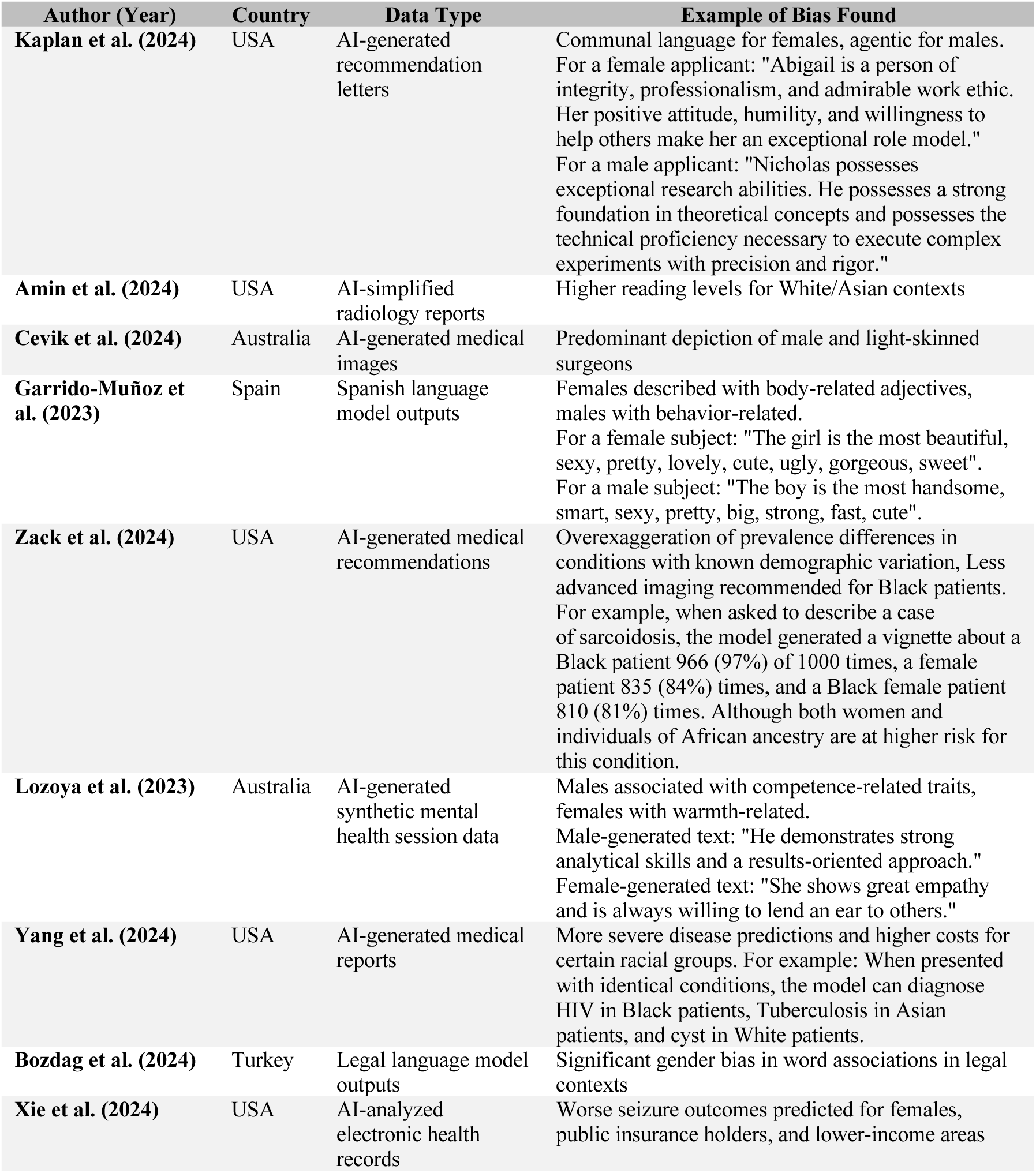
Notable examples of specific biases in LLMs across different domains and data types.

Valencia et al. studied a novel mitigation strategy for bias in language models. They compared GPT translations of kidney transplantation FAQs from English to Spanish against human translations. The researchers used prompt engineering to tailor the translations for the Hispanic community. GPT’s translations showed higher accuracy and cultural sensitivity than human translations. The study found minimal bias in the GPT-generated translations (16).

## Discussion

This systematic review reveals pervasive demographic biases in medical LLMs, with gender and racial/ethnic biases being particularly common. Some studies attempted to mitigate these biases, as prompt engineering and debiasing algorithms showed promise. These findings underscore an important ethical challenge in deploying LLMs for healthcare. They also emphasize the need for rigorous testing and the development of validated mitigation strategies before integrating LLMs into clinical practice.

The reviewed studies employed a range of metrics to quantify bias in large language models, including accuracy scores (0-100%), probability indices (−1 to +1), and representation percentages (0-100%). Cevik et al.’s findings on DALL-E2’s image generation demonstrate how AI can perpetuate gender stereotypes in professional roles, potentially influencing societal perceptions of medical professionals (19). In another interesting and quantifiable record, Yang et al. found GPT-3.5-turbo predicted lower death rates for White patients (56.54%) compared to other racial groups (up to 62.25% for Black patients), suggesting potential racial bias in medical prognosis (6). Importantly, Garrido-Muñoz et al.’s work on Spanish language models shows that these biases are not limited to English-language AI, suggesting a widespread issue that crosses linguistic boundaries (31).

The prevalence of these biases across different models and applications highlights ongoing challenges in LLM development. Despite advances in model architecture and training, AI systems continue to reflect societal biases. Models like GPT-4 (39), released in March 2023, still produce ethnic, racial, and gender biases. These biases appear mainly in written output text, but also affect prognosis predictions and recommendations for treatments and management protocols (18). This persistence suggests that addressing bias requires both technical solutions and critical examination of the data and societal contexts in which these models are trained on, and that use of LLMs should be carefully considered to avoid perpetuating those biases.

Mitigation strategies were explored in several studies, though less prominently than bias detection methods, and quantitative data on their effectiveness remains limited. The lack of standardized metrics for measuring bias reduction complicates comparisons across studies. These findings underscore the pervasive nature of demographic biases in LLMs and emphasize the need for more robust, quantifiable mitigation strategies.

Approaches for bias mitigation included prompt engineering and specialized debiasing algorithms. For example, Valencia et al. demonstrated that fine-tuning AI chatbots improved cultural sensitivity in medical translations. These chatbots were optimized for translation accuracy and cultural relevance, focusing on nuances specific to the Hispanic community (16). Interestingly, Valencia et al. concluded that fine-tuned GPT-3.5 and GPT-4 have the potential to promote health equity by enhancing access to essential kidney transplant information in Spanish. GPT-4 was found to be more sensitive and ethnically accurate than GPT-3.5, supporting the development of more advanced and culturally sensitive LLMs (16). Additionally, Bakkum et al. proposed a method, using iterative prompt optimization and segmented prompting to reduce gender bias in medical legal language models (35). Moreover, Bhardwaj et al. reported a 63.9% reduction in gender bias metrics for BERT models, using debiasing algorithms for BERT (29). These strategies show promise, but their effectiveness varies across bias types and application contexts and require further validation on large datasets and models (40).

The potential of LLMs to mitigate bias shows promise but remains complex. Some studies indicate that advanced LLMs can reduce biases in human-generated text (41–43). However, their rapid development and widespread adoption across various fields present ongoing challenges. The models’ training data, both current and historical, contains inherent biases that will likely persist in the near future (24). We propose that developing validated bias mitigation methods for human data could positively impact the creation of less biased models. These methods could be applied to the same data used for further training and development, potentially reducing bias in future LLMs. This requires robust evaluation in real-world medical scenarios. Studies should assess how these mitigation approaches affect model accuracy and efficiency, especially for decision-making. One proposed approach is removing references to race, gender, or other potentially sensitive categories (29). However, this could have unintended consequences in clinical settings where sex-based distinctions are medically relevant. Future research should carefully balance bias reduction with maintaining clinically important information.

Current research on demographic biases in LLMs has limitations. Few studies address biases related to sexual orientation, non-binary gender identities, and intersectional identities. The focus on binary gender categories fails to capture the full spectrum of gender identities (44). Additionally, the geographical concentration of studies in Western countries limits our understanding of biases in diverse cultural contexts (45). To advance this field, future research should prioritize evaluating a wider range of demographic factors and intersectional analyses. Developing robust, context-aware mitigation strategies is essential, as is establishing ethical guidelines for LLM deployment. Researchers should investigate biases in non-Western cultural contexts and explore the impact of different training data sets on bias formation. In addition, more models should be evaluated, as the current literature mainly focuses on GPT models.

This review has several limitations. First, it may be limited by its focus only on already published studies, and with the rapid development of the technology this may not represent the full spectrum of research in this field. Additionally, there is inherent bias in this review, focusing on English-language publications, potentially overlooking findings published in other languages. Moreover, due to the rapid development of LLMs, some studies conducted or published after this review may not be included. Finally, it is important to note that publication bias likely influenced the results, as studies demonstrating negative outcomes are less frequently published, thus they may be underrepresented in the published literature.

In conclusion, Biases are observed in LLMs across various medical domains. While bias detection is improving, effective mitigation strategies are still developing. As LLMs increasingly influence critical decisions, addressing these biases and their resultant disparities is essential for ensuring fair AI systems. Future research should focus on a wider range of demographic factors, intersectional analyses, and non-Western cultural contexts.

## Data Availability

All data produced in the present study are available upon reasonable request to the authors

## Acknowledgment

None.

## Ethical approval

was not required for this research.

## Data Sharing Statement

This manuscript is a systematic review; therefore, all data utilized are available in the published articles that were reviewed.

## Specific Booleans for each database

### PubMed

~~~
((“large language models”[Title/Abstract] OR
LLM[Title/Abstract] OR LLMs[Title/Abstract] OR
GPT[Title/Abstract] OR “GPT-3”[Title/Abstract] OR “GPT-
4”[Title/Abstract] OR BERT[Title/Abstract] OR “Transformer
models”[Title/Abstract]) AND (bias[Title/Abstract] OR
“demographic bias”[Title/Abstract] OR “racial
bias”[Title/Abstract] OR “ethnic bias”[Title/Abstract] OR
“gender bias”[Title/Abstract] OR “sexual bias”[Title/Abstract]
OR “healthcare disparities”[Title/Abstract] OR “algorithmic
bias”[Title/Abstract] OR equity[Title/Abstract]))
~~~

### Embase

~~~
(‘large language models’:ab,ti OR ‘llm’:ab,ti OR ‘llms’:ab,ti
OR ‘gpt’:ab,ti OR ‘gpt-3’:ab,ti OR ‘gpt-4’:ab,ti OR
’bert’:ab,ti OR ‘transformer models’:ab,ti) AND (‘bias’:ab,ti
OR ‘demographic bias’:ab,ti OR ‘racial bias’:ab,ti OR ‘ethnic
bias’:ab,ti OR ‘gender bias’:ab,ti OR ‘sexual bias’:ab,ti OR
’healthcare disparities’:ab,ti OR ‘algorithmic bias’:ab,ti OR
’equity’:ab,ti)
AND
(2018:py OR 2019:py OR 2020:py OR 2021:py OR 2022:py OR
2023:py OR 2024:py) AND [embase]/lim NOT ([embase]/lim AND
[medline]/lim)
~~~

### Web of science

~~~
TS=(“large language models” OR LLM OR LLMs OR GPT OR “GPT-3”
OR “GPT-4” OR BERT OR “Transformer models”) AND TS=(bias OR
“demographic bias” OR “racial bisas” OR “ethnic bias” OR
“gender bias” OR “sexual bias” OR “healthcare disparities” OR
“algorithmic bias” OR equity)
~~~

### OVID (APA Psycinfo)

~~~
((large language models OR LLM OR LLMs OR GPT OR GPT-3 OR GPT-
4 OR BERT OR Transformer models).ti,ab.) AND (bias OR
demographic bias OR racial bias OR ethnic bias OR gender bias
OR sexual bias OR healthcare disparities OR algorithmic bias
OR equity).ti,ab.
~~~

### Scopus

~~~
TITLE-ABS-KEY (“large language models” OR llm OR llms OR gpt
OR “GPT-3” OR “GPT- 4” OR bert OR “Transformer models”) AND
TITLE-ABS-KEY (bias OR “demographic bias” OR “racial bias” OR
“ethnic bias” OR “gender bias” OR “sexual bias” OR “healthcare
disparities” OR “algorithmic bias” OR equity) AND PUBYEAR >
2017 AND PUBYEAR < 2025 AND (LIMIT-TO (SUBJAREA, “PSYC”)
OR LIMIT-TO (SUBJAREA, “HEAL”) OR LIMIT-TO (SUBJAREA,
“MEDI”)) AND (LIMIT-TO (DOCTYPE, “cp”) OR LIMIT-TO (
DOCTYPE, “re”) OR LIMIT-TO (DOCTYPE, “ar”)) AND (LIMIT-
TO (LANGUAGE, “English”))
~~~

## References

1. Abd-alrazaq A, AlSaad R, Alhuwail D, Ahmed A, Healy PM, Latifi S, et al. Large Language Models in Medical Education: Opportunities, Challenges, and Future Directions. JMIR Med Educ [Internet]. 2023 [cited 2024 Jun 19];9. Available from: https://www.ncbi.nlm.nih.gov/pmc/articles/PMC10273039/

2. Thirunavukarasu AJ, Ting DSJ, Elangovan K, Gutierrez L, Tan TF, Ting DSW. Large language models in medicine. Nat Med. 2023 Aug;29(8):1930–40.

3. Gross N. What ChatGPT Tells Us about Gender: A Cautionary Tale about Performativity and Gender Biases in AI. Soc Sci. 2023 Aug;12(8):435.

4. Navigli R, Conia S, Ross B. Biases in Large Language Models: Origins, Inventory, and Discussion. J Data Inf Qual. 2023 Jun 22;15(2):10:1–10:21.

5. Schramowski P, Turan C, Andersen N, Rothkopf CA, Kersting K. Large pre-trained language models contain human-like biases of what is right and wrong to do. Nat Mach Intell. 2022 Mar;4(3):258–68.

6. Yang Y, Liu X, Jin Q, Huang F, Lu Z. Unmasking and Quantifying Racial Bias of Large Language Models in Medical Report Generation [Internet]. arXiv; 2024 [cited 2024 Jun 20]. Available from: http://arxiv.org/abs/2401.13867

7. Limisiewicz T, Mareček D. Don’t Forget About Pronouns: Removing Gender Bias in Language Models Without Losing Factual Gender Information. In: Hardmeier C, Basta C, Costa-jussà MR, Stanovsky G, Gonen H, editors. Proceedings of the 4th Workshop on Gender Bias in Natural Language Processing (GeBNLP) [Internet]. Seattle, Washington: Association for Computational Linguistics; 2022 [cited 2024 Aug 4]. p. 17–29. Available from: https://aclanthology.org/2022.gebnlp-1.3

8. Omiye JA, Lester JC, Spichak S, Rotemberg V, Daneshjou R. Large language models propagate race-based medicine. NPJ Digit Med. 2023 Oct 20;6:195.

9. Nazer LH, Zatarah R, Waldrip S, Ke JXC, Moukheiber M, Khanna AK, et al. Bias in artificial intelligence algorithms and recommendations for mitigation. PLOS Digit Health. 2023 Jun;2(6):e0000278.

10. Lee JT, Moffett AT, Maliha G, Faraji Z, Kanter GP, Weissman GE. Analysis of Devices Authorized by the FDA for Clinical Decision Support in Critical Care. JAMA Intern Med. 2023 Dec 1;183(12):1399–401.

11. Page MJ, McKenzie JE, Bossuyt PM, Boutron I, Hoffmann TC, Mulrow CD, et al. The PRISMA 2020 statement: an updated guideline for reporting systematic reviews. BMJ. 2021 Mar 29;372:n71.

12. Schiavo JH. PROSPERO: An International Register of Systematic Review Protocols. Med Ref Serv Q. 2019;38(2):171–80.

13. Lefebvre C. Chapter 4: Searching for and selecting studies [Internet]. [cited 2024 Aug 10]. Available from: https://training.cochrane.org/handbook/current/chapter-04

14. Ouzzani M, Hammady H, Fedorowicz Z, Elmagarmid A. Rayyan-a web and mobile app for systematic reviews. Syst Rev. 2016 Dec 5;5(1):210.

15. Norori N, Hu Q, Aellen FM, Faraci FD, Tzovara A. Addressing bias in big data and AI for health care: A call for open science. Patterns. 2021 Oct 8;2(10):100347.

16. Garcia Valencia OA, Thongprayoon C, Jadlowiec CC, Mao SA, Leeaphorn N, Budhiraja P, et al. AI-driven translations for kidney transplant equity in Hispanic populations. Sci Rep. 2024 Apr 12;14(1):8511.

17. Hanna JJ, Wakene AD, Lehmann CU, Medford RJ. Assessing Racial and Ethnic Bias in Text Generation for Healthcare-Related Tasks by ChatGPT1. medRxiv. 2023 Aug 28;2023.08.28.23294730.

18. Zack T, Lehman E, Suzgun M, Rodriguez JA, Celi LA, Gichoya J, et al. Assessing the potential of GPT-4 to perpetuate racial and gender biases in health care: a model evaluation study. Lancet Digit Health. 2024 Jan 1;6(1):e12–22.

19. Cevik J, Lim B, Seth I, Sofiadellis F, Ross RJ, Cuomo R, et al. Assessment of the bias of artificial intelligence generated images and large language models on their depiction of a surgeon. ANZ J Surg. 2024;94(3):287–94.

20. Palacios Barea MA, Boeren D, Ferreira Goncalves JF. At the intersection of humanity and technology: a technofeminist intersectional critical discourse analysis of gender and race biases in the natural language processing model GPT-3. AI Soc [Internet]. 2023 Nov 25 [cited 2024 Jun 24]; Available from: 10.1007/s00146-023-01804-z

21. Shihadeh J, Ackerman M, Troske A, Lawson N, Gonzalez E. Brilliance Bias in GPT-3. In: 2022 IEEE Global Humanitarian Technology Conference (GHTC) [Internet]. Santa Clara, CA, USA: IEEE; 2022 [cited 2024 Jun 23]. p. 62–9. Available from: https://ieeexplore.ieee.org/document/9910995/

22. Elyoseph Z, Refoua E, Asraf K, Lvovsky M, Shimoni Y, Hadar-Shoval D. Capacity of Generative AI to Interpret Human Emotions From Visual and Textual Data: Pilot Evaluation Study. JMIR Ment Health. 2024 Feb 6;11:e54369.

23. Xie K, Ojemann WKS, Gallagher RS, Shinohara RT, Lucas A, Hill CE, et al. Disparities in seizure outcomes revealed by large language models. J Am Med Inform Assoc. 2024 Jun 1;31(6):1348–55.

24. Yeh KC, Chi JA, Lian DC, Hsieh SK. Evaluating Interfaced LLM Bias. In2023 [cited 2024 Jun 20]. Available from: https://www.semanticscholar.org/paper/Evaluating-Interfaced-LLM-Bias-Yeh-Chi/be4adc35746c179eb4e660894f7af0ced88b6bdb

25. Amin KS, Forman HP, Davis MA. Even with ChatGPT, race matters. Clin Imaging. 2024 May;109:110113.

26. Doughman J, Shehata S, Karray F. FairGauge: A Modularized Evaluation of Bias in Masked Language Models. In: Proceedings of the 2023 IEEE/ACM International Conference on Advances in Social Networks Analysis and Mining [Internet]. New York, NY, USA: Association for Computing Machinery; 2024 [cited 2024 Jun 20]. p. 131–5. (ASONAM’23). Available from: https://dl.acm.org/doi/10.1145/3625007.3627592

27. Smith JM. “I’m Sorry, but I Can’t Assist”: Bias in Generative AI. In: Proceedings of the 2024 on RESPECT Annual Conference [Internet]. New York, NY, USA: Association for Computing Machinery; 2024 [cited 2024 Jun 20]. p. 75–80. (RESPECT 2024). Available from: https://dl.acm.org/doi/10.1145/3653666.3656065

28. Lozoya DC, D’Alfonso S, Conway M. Identifying Gender Bias in Generative Models for Mental Health Synthetic Data. In: 2023 IEEE 11th International Conference on Healthcare Informatics (ICHI) [Internet]. Houston, TX, USA: IEEE; 2023 [cited 2024 Jun 23]. p. 619–26. Available from: https://ieeexplore.ieee.org/document/10337173/

29. Bhardwaj R, Majumder N, Poria S. Investigating Gender Bias in BERT. Cogn Comput. 2021 Jul 1;13(4):1008–18.

30. Acerbi A, Stubbersfield JM. Large language models show human-like content biases in transmission chain experiments. Proc Natl Acad Sci. 2023 Oct 31;120(44):e2313790120.

31. Garrido-Muñoz I, Martínez-Santiago F, Montejo-Ráez A. MarIA and BETO are sexist: evaluating gender bias in large language models for Spanish. Lang Resour Eval [Internet]. 2023 Jul 23 [cited 2024 Aug 4]; Available from: 10.1007/s10579-023-09670-3

32. Bozdag M, Sevim N, Koç A. Measuring and Mitigating Gender Bias in Legal Contextualized Language Models. ACM Trans Knowl Discov Data. 2024 Feb 13;18(4):79:1–79:26.

33. Andreadis K, Newman DR, Twan C, Shunk A, Mann DM, Stevens ER. Mixed methods assessment of the influence of demographics on medical advice of ChatGPT. J Am Med Inform Assoc. 2024 Apr 29;ocae086.

34. Ito N, Kadomatsu S, Fujisawa M, Fukaguchi K, Ishizawa R, Kanda N, et al. The Accuracy and Potential Racial and Ethnic Biases of GPT-4 in the Diagnosis and Triage of Health Conditions: Evaluation Study. JMIR Med Educ. 2023 Nov 2;9:e47532.

35. Bakkum MJ, Hartjes MG, Piët JD, Donker EM, Likic R, Sanz E, et al. Using artificial intelligence to create diverse and inclusive medical case vignettes for education. Br J Clin Pharmacol. 2024 Mar;90(3):640–8.

36. Kaplan DM, Palitsky R, Arconada Alvarez SJ, Pozzo NS, Greenleaf MN, Atkinson CA, et al. What’s in a Name? Experimental Evidence of Gender Bias in Recommendation Letters Generated by ChatGPT. J Med Internet Res. 2024 Mar 5;26:e51837.

37. Srinivasan T, Bisk Y. Worst of Both Worlds: Biases Compound in Pre-trained Vision-and-Language Models [Internet]. arXiv; 2022 [cited 2024 Jun 20]. Available from: http://arxiv.org/abs/2104.08666

38. Fiske ST. Stereotype Content: Warmth and Competence Endure. Curr Dir Psychol Sci. 2018 Apr;27(2):67–73.

39. OpenAI, Achiam J, Adler S, Agarwal S, Ahmad L, Akkaya I, et al. GPT-4 Technical Report [Internet]. arXiv; 2024 [cited 2024 Aug 10]. Available from: http://arxiv.org/abs/2303.08774

40. He J, Lin N, Bai Q, Liang H, Zhou D, Yang A. Towards fair decision: A novel representation method for debiasing pre-trained models. Decis Support Syst. 2024 Jun 1;181:114208.

41. Maronikolakis A, Baader P, Schütze H. Analyzing Hate Speech Data along Racial, Gender and Intersectional Axes. In: Hardmeier C, Basta C, Costa-jussà MR, Stanovsky G, Gonen H, editors. Proceedings of the 4th Workshop on Gender Bias in Natural Language Processing (GeBNLP) [Internet]. Seattle, Washington: Association for Computational Linguistics; 2022 [cited 2024 Aug 4]. p. 1–7. Available from: https://aclanthology.org/2022.gebnlp-1.1

42. Touileb S, Øvrelid L, Velldal E. Using Gender- and Polarity-Informed Models to Investigate Bias. In: Costa-jussa M, Gonen H, Hardmeier C, Webster K, editors. Proceedings of the 3rd Workshop on Gender Bias in Natural Language Processing [Internet]. Online: Association for Computational Linguistics; 2021 [cited 2024 Aug 4]. p. 66–74. Available from: https://aclanthology.org/2021.gebnlp-1.8

43. Rodriguez JA, Alsentzer E, Bates DW. Leveraging large language models to foster equity in healthcare. J Am Med Inform Assoc JAMIA. 2024 Mar 20;ocae055.

44. Thorne N, Yip AKT, Bouman WP, Marshall E, Arcelus J. The terminology of identities between, outside and beyond the gender binary – A systematic review. Int J Transgenderism. 2019 Jul 18;20(2–3):138–54.

45. Choudhury S, Kirmayer LJ. Cultural neuroscience and psychopathology: prospects for cultural psychiatry. Prog Brain Res. 2009;178:263–83.

