## Supplementary materials for "Evaluating and Addressing Demographic Disparities in Medical Large Language Models: A Systematic Review"

#### Index

**_Boolean strings for each database_** ........ **_2_**

**_Screening and Inclusion Process_** ........ **_4_**

**_Data Extraction Process_** ........ **_5_**

**_Quality assessment_** ........ **_7_**

***Table S1****: the Critical Appraisal Checklist for Analytical Cross-Sectional Studies.* ........ *7*

***Table S2****: the Critical Appraisal Checklist for Diagnostic Test Accuracy Studies.* ........ *8*

**_Other supplemental figures and tables_** ........ **_9_**

### Boolean strings for each database

#### PubMed

(("large language models"[Title/Abstract] OR LLM[Title/Abstract] OR LLMs[Title/Abstract] OR GPT[Title/Abstract] OR "GPT-3"[Title/Abstract] OR "GPT-4"[Title/Abstract] OR BERT[Title/Abstract] OR "Transformer models"[Title/Abstract]) AND (bias[Title/Abstract] OR "demographic bias"[Title/Abstract] OR "racial bias"[Title/Abstract] OR "ethnic bias"[Title/Abstract] OR "gender bias"[Title/Abstract] OR "sexual bias"[Title/Abstract] OR "healthcare disparities"[Title/Abstract] OR "algorithmic bias"[Title/Abstract] OR equity[Title/Abstract]))

#### Embase

('large language models':ab,ti OR 'llm':ab,ti OR 'llms':ab,ti OR 'gpt':ab,ti OR 'gpt-3':ab,ti OR 'gpt-4':ab,ti OR 'bert':ab,ti OR 'transformer models':ab,ti) AND ('bias':ab,ti OR 'demographic bias':ab,ti OR 'racial bias':ab,ti OR 'ethnic bias':ab,ti OR 'gender bias':ab,ti OR 'sexual bias':ab,ti OR 'healthcare disparities':ab,ti OR 'algorithmic bias':ab,ti OR 'equity':ab,ti)

AND

(2018:py OR 2019:py OR 2020:py OR 2021:py OR 2022:py OR 2023:py OR 2024:py) AND [embase]/lim NOT ([embase]/lim AND [medline]/lim)

#### Web of science

TS=("large language models" OR LLM OR LLMs OR GPT OR "GPT-3" OR "GPT-4" OR BERT OR "Transformer models") AND TS=(bias OR "demographic bias" OR "racial bisas" OR "ethnic bias" OR "gender bias" OR "sexual bias" OR "healthcare disparities" OR "algorithmic bias" OR equity)

#### OVID (APA Psycinfo)

((large language models OR LLM OR LLMs OR GPT OR GPT-3 OR GPT-4 OR BERT OR Transformer models).ti,ab.) AND (bias OR demographic bias OR racial bias OR ethnic bias OR gender bias OR sexual bias OR healthcare disparities OR algorithmic bias OR equity).ti,ab.

#### Scopus

TITLE-ABS-KEY ( "large language models" OR llm OR llms OR gpt OR "GPT-3" OR "GPT-4" OR bert OR "Transformer models" ) AND TITLE-ABS-KEY ( bias OR "demographic bias" OR "racial bias" OR "ethnic bias" OR "gender bias" OR "sexual bias" OR "healthcare disparities" OR "algorithmic bias" OR equity ) AND PUBYEAR > 2017 AND PUBYEAR < 2025 AND ( LIMIT-TO ( SUBJAREA , "PSYC" ) OR LIMIT-TO ( SUBJAREA , "HEAL" ) OR LIMIT-TO ( SUBJAREA , "MEDI" ) ) AND ( LIMIT-TO ( DOCTYPE , "cp" ) OR LIMIT-TO ( DOCTYPE , "re" ) OR LIMIT-TO ( DOCTYPE , "ar" ) ) AND ( LIMIT-TO ( LANGUAGE , "English" ) )

#### Screening and Inclusion Process

Two reviewers (MO and EK) independently screened titles and abstracts of all identified records. The reviewers used the following inclusion criteria:

- Peer-reviewed study
- Evaluated demographic biases in large language models (LLMs)
- Applied to medical or healthcare tasks

Demographic bias was defined as systematic variation in model outputs based on characteristics such as gender, race, ethnicity, age, or socioeconomic status.

The reviewers excluded:

- Studies of non-LLM artificial intelligence models
- Studies focusing solely on model performance without addressing bias
- Non-peer-reviewed materials (e.g. preprints, conference abstracts)

After independent screening, the reviewers compared their decisions. Any disagreements were resolved through discussion. For records where agreement could not be reached, a third reviewer arbitrated.

Full-text articles were obtained for all records deemed potentially eligible after title/abstract screening. The same two reviewers then independently assessed the full-text articles against the inclusion/exclusion criteria. Again, disagreements were resolved through discussion or arbitration by the third reviewer if needed.

#### Data Extraction Process

Two reviewers (Reviewer 1 and Reviewer 2) independently extracted data from each included study using a standardized form. The form captured the following information:

1. Study Characteristics

- Author: Last name of first author followed by "et al."
- Year of Publication
- Country where research was conducted
- Study design and methodology specifics for quality assessment
- Objective/Aims: Concise summary focused on evaluating bias, equity, or diversity in LLM use
- Sample Size

2. Data and Model Details

- Type of data used to evaluate LLM performance
- LLM(s) evaluated (e.g. GPT-3.5, GPT-4)

3. Bias Assessment

- Type(s) of bias studied (e.g. racial, ethnic, gender, sexual orientation, socioeconomic)
- Bias detection methods and tools
- Bias mitigation strategies (if used) and their effects

4. Key Findings

- Summary of main results on demographic biases identified
- Performance metrics (e.g. F1 score, accuracy, precision, recall)
- Percentage of cases exhibiting bias

5. Implications and Limitations

- Authors' conclusions on LLM demographic biases
- Proposed bias mitigation strategies
- Study limitations (stated or inferred)

6. Additional Information

- Interesting findings or specific bias examples

The reviewers independently extracted data into the form for each study. They then compared their extractions and resolved any discrepancies through discussion or consultation with a third reviewer.

To ensure consistency, the reviewers first piloted the form on 3 included studies, refining the extraction process before proceeding with all studies.

For studies with missing information, the reviewers made reasonable inferences where possible, clearly marking these as inferred. If critical information was missing, the reviewers contacted study authors for clarification.

The extracted data was compiled into summary tables for analysis. One reviewer entered the final agreed data into the tables, with a second reviewer checking for accuracy.

### Quality assessment

#### **Table S1**: the Critical Appraisal Checklist for Analytical Cross-Sectional Studies.

| *Study* | *D1* | *D2* | *D3* | *D4* | *D5* | *D6* | *D7* | *D8* |
|---|---|---|---|---|---|---|---|---|
| *Valencia et al.* | Yes | Yes | Yes | Yes | Unclear | Unclear | Yes | Yes |
| *Elyoseph et al.* | Yes | Yes | Yes | Yes | Unclear | No | Yes | Yes |
| *Cevik et al.* | Yes | Yes | Yes | Yes | No | No | Yes | No |
| *Kaplan et al.* | Yes | Yes | Yes | Yes | Yes | Yes | Yes | Yes |
| *Amin et al.* | Yes | Yes | Yes | Yes | Yes | Yes | Yes | Yes |
| *Bakkum et al.* | Yes | Yes | Yes | Yes | Yes | Yes | Yes | No |
| *Smith et al.* | Yes | Yes | Yes | Yes | Yes | Yes | Yes | Yes |
| *Yang et al.* | Yes | Yes | Yes | Yes | Unclear | No | Yes | Yes |
| *Lozoya et al.* | Yes | Yes | Yes | Yes | No | No | Yes | Yes |
| *Shihadeh et al.* | Yes | Yes | Yes | Yes | Yes | No | No | Yes |
| *Yeh et al.* | Yes | Yes | Yes | Yes | Unclear | No | Yes | No |
| *Doughman et al.* | Yes | Yes | Yes | Yes | No | No | Yes | Yes |
| *Srinivasan et al.* | Yes | Yes | Yes | Yes | Yes | Yes | Yes | Yes |
| *Zack et al.* | NA | Yes | Yes | Yes | Yes | Yes | Yes | Yes |
| *Hanna et al.* | Yes | Yes | Yes | Yes | Yes | Yes | Yes | Yes |
| *Acerbi et al.* | Yes | Yes | Yes | Yes | Yes | Yes | Yes | Yes |
| *Bhardwaj et al.* | Yes | Yes | Yes | Yes | Yes | Yes | Yes | Yes |
| *Palacios Barea et al.* | Yes | yes | Yes | Yes | Yes | No | Yes | NA |
| *Garrido-Muñoz et al.* | Yes | yes | Yes | Yes | Yes | No | Yes | Yes |
| *Bozdag et al.* | Yes | Yes | Yes | Yes | Yes | Yes | Yes | Yes |
| *Gross et al.* | NA | NA | Yes | Yes | No | No | Yes | NA |

**Abbreviations**:

- D1: Were the criteria for inclusion in the sample clearly defined?
- D2: Were the study subjects and the setting described in detail?
- D3: Was the exposure measured in a valid and reliable way?
- D4: Were objective, standard criteria used for measurement of the condition?
- D5: Were confounding factors identified?
- D6: Were strategies to deal with confounding factors stated?
- D7: Were the outcomes measured in a valid and reliable way?
- D8: Was appropriate statistical analysis used?
- NA: Not Applicable

#### **Table S2**: the Critical Appraisal Checklist for Diagnostic Test Accuracy Studies.

| ***Study*** | *D1* | *D2* | *D3* | *D4* | *D5* | *D6* | *D7* | *D8* | *D9* | *D10* |
|---|---|---|---|---|---|---|---|---|---|---|
| *Ito et al.* | NA | Yes | Yes | Yes | Yes | Yes | Yes | NA | Yes | Yes |
| *Xie et al.* | No | Yes | Yes | Yes | NA | Yes | Unclear | NA | Yes | Yes |
| *Andreadis et al.* | NA | Yes | Yes | NA | NA | Yes | Yes | NA | Yes | Yes |

**Abbreviations**:

- D1: Was a consecutive or random sample of patients enrolled?
- D2: Was a case-control design avoided?
- D3: Did the study avoid inappropriate exclusions?
- D4: Were the index test results interpreted without knowledge of the results of the reference standard?
- D5: If a threshold was used, was it pre-specified?
- D6: Is the reference standard likely to correctly classify the target condition?
- D7: Were the reference standard results interpreted without knowledge of the results of the index test?
- D8: Was there an appropriate interval between index test and reference standard?
- D9: Did all patients receive the same reference standard?
- D10: Were all patients included in the analysis?
- NA: Not Applicable

### Other supplemental figures and tables

**Table S3:** Detailed methodological and quantitative analysis of the included studies.

| Author et al. | Model | Type of bias | Sample size | Type of data | Bias Detection Methods | Numeric results | Study limitations |
|---|---|---|---|---|---|---|---|
| **Elyoseph et al.** | GPT-4, Google Bard | Gender | 56 items | Visual and textual data | Statistical analysis | nearly equal distribution of errors across male (9) and female (10) stimuli in RMET | Limited to specific emotion recognition tasks |
| **Kaplan et al.** | GPT-3.5 | Gender | 1400 letters | AI-generated text | LIWC analysis | Significant differences in language use ($p < 0.05$) | Focus on binary gender, limited name set |
| **Bakkum et al.** | GPT-3.5 | Gender | Not reported | AI-generated case vignettes | Not specified | Not numerically evaluated | Limited to medical case generation |
| **Bhardwaj et al.** | BERT | Gender | 8,400 samples | Template-based sentences | MLP regressors, equity evaluation | Not numerically evaluated | Limited to binary gender attributes |
| **Shihadeh et al.** | GPT-3, InstructGPT | Gender | 3200 generations | AI-generated text | Template-based approach | Not numerically evaluated | Focus on brilliance bias only |
| **Garrido-Muñoz et al.** | Various Spanish LLMs | Gender | 20 templates | Masked language task | Probability and rank-based metrics | Not numerically evaluated | Limited to Spanish language models |
| **Srinivasan et al.** | VL-BERT | Gender | 12 images per entity pair | Image-text pairs | Template-based masked language modeling | Not numerically evaluated | Limited to binary gender classification |
| **Bozdag et al.** | LegalBERT-Small | Gender | 3,032 court cases | Legal documents | Template-based approach | Comparable μ-F1 and m-F1 scores after debiasing | Specific to legal domain |
| **Gross et al.** | GPT-4 | Gender | Not applicable | AI-generated responses | Qualitative analysis | Not reported | Conceptual nature, lack of empirical data |
| **Lozoya et al.** | GPT-3 | Gender | 1,000 text documents | Synthetic text data | LIWC-22, ccLDA | Significant differences in trait associations ($p < 0.05$) | Context-specific to mental health therapy |
| **Cevik et al.** | GPT-3.5, BARD | Gender, racial | 24 descriptions, 64 images | AI-generated images and text | Analysis of generated descriptions and images | Not numerically evaluated | Limited group of evaluators |
| **Palacios Barea et al.** | GPT-3 | Gender, racial | 56 unique prompts | Text completions | Critical Discourse Analysis | Not numerically evaluated | Stochastic nature of outputs, researcher bias |
| **Acerbi et al.** | GPT-3 | Gender, social, threat-related | Not reported | AI-generated text | Transmission chain method | Not numerically evaluated | Limited to specific content biases |
| **Doughman et al.** | BERT, DistilBERT | Gender, racial, class, religious | 23,736 sentences | Synthetically generated prejudiced sentences | Prejudice score combining probability and top-k index | Sexism had highest match rate (BERT: 24%, DistilBERT: 16%) | Use of synthetic data may not reflect real-world language |

| | | | | | | | |
|---|---|---|---|---|---|---|---|
| **Smith et al.** | GPT-3.5, Claude AI | Racial, ethnic | 100 hypothetical students | AI-generated recommendations | Algorithm audit approach | Significant differences in recommendations ($p < 0.05$) | Limited to specific academic context |
| **Amin et al.** | GPT-3.5, GPT-4 | Racial, ethnic | 750 reports | Radiology reports | Readability scores | Significant differences in reading grade levels ($p < 0.05$) | Focus only on readability scores |
| **Yang et al.** | GPT-3.5-turbo, GPT-4 | Racial | 383 patient profiles | AI-generated medical reports | Qualitative and quantitative analyses | Lower death rate prediction for White patients (56.54% vs. others) | Limited to specific medical scenarios |
| **Hanna et al.** | GPT-3.5 | Racial, ethnic | 100 encounters | Health record encounters | Sentiment analysis, NER, readability scores | No significant differences across races/ethnicities ($p > 0.05$) | Reliance on specific linguistic measures |
| **Ito et al.** | GPT-4 | Racial, ethnic | 45 | Clinical vignettes | Comparison with physician performance | 97.8% diagnostic accuracy, no significant differences across groups | Limited set of clinical vignettes |
| **Xie et al.** | Clinical_BERT | Racial, ethnic, gender, socioeconomic | 84,675 clinic visits | Electronic Health Records | Accuracy of model classifications, regression models | Significant disparities in outcomes across demographic groups ($p < 0.05$) | Potential biases in human annotations |
| **Zack et al.** | GPT-4 | Racial, ethnic, gender | 1000 | Clinical vignettes | Statistical tests for significance between groups | Significant differences in diagnosis and treatment recommendations by race and gender ($p < 0.05$) | Limited to specific medical context |
| **Andreadis et al.** | GPT-4 | Racial, ethnic, age, sex | 540 | Clinical vignettes | Analysis of diagnostic correctness and generated text | No significant difference in diagnostic accuracy, but age bias in recommendations | May not reflect all patient interactions |
| **Valencia et al.** | GPT-3.5, GPT-4.0 | Cultural, linguistic | 54 questions | Translated medical FAQs | Evaluation by native speakers using 1-5 rubric | High linguistic accuracy (GPT-3.5: 4.89 ± 0.31, GPT-4.0: 4.94 ± 0.23) | Narrow focus on specific medical context |
| **Yeh et al.** | GPT-3.5 | Age, disability, socioeconomic, sexual orientation | Not reported | Content and comments from online board | Sentiment analysis, bias QA dataset | Not numerically evaluated | Reliance on specific dataset, potential keyword biases |

***Abbreviations**: LLMs: Large Language Models | LIWC: Linguistic Inquiry and Word Count | MLP: Multilayer Perceptron | NER: Named Entity Recognition | BERT: Bidirectional Encoder Representations from Transformers | GPT: Generative Pre-trained Transformer | AI: Artificial Intelligence | ccLDA: cross-collection Latent Dirichlet Allocation | FAQ: Frequently Asked Questions | QA: Question Answering

**Table S4**: Notable examples of specific biases in LLMs across different domains and data types.

| Author (Year) | Country | Data Type | Example of Bias Found |
|---|---|---|---|
| **Kaplan et al. (2024)** | USA | AI-generated recommendation letters | Communal language for females, agentic for males. For a female applicant: "Abigail is a person of integrity, professionalism, and admirable work ethic. Her positive attitude, humility, and willingness to help others make her an exceptional role model." For a male applicant: "Nicholas possesses exceptional research abilities. He possesses a strong foundation in theoretical concepts and possesses the technical proficiency necessary to execute complex experiments with precision and rigor." |
| **Amin et al. (2024)** | USA | AI-simplified radiology reports | Higher reading levels for White/Asian contexts |
| **Cevik et al. (2024)** | Australia | AI-generated medical images | Predominant depiction of male and light-skinned surgeons |
| **Garrido-Muñoz et al. (2023)** | Spain | Spanish language model outputs | Females described with body-related adjectives, males with behavior-related. For a female subject: "The girl is the most beautiful, sexy, pretty, lovely, cute, ugly, gorgeous, sweet". For a male subject: "The boy is the most handsome, smart, sexy, pretty, big, strong, fast, cute". |
| **Zack et al. (2024)** | USA | AI-generated medical recommendations | Overexaggeration of prevalence differences in conditions with known demographic variation, Less advanced imaging recommended for Black patients. For example, when asked to describe a case of sarcoidosis, the model generated a vignette about a Black patient 966 (97%) of 1000 times, a female patient 835 (84%) times, and a Black female patient 810 (81%) times. Although both women and individuals of African ancestry are at higher risk for this condition. |
| **Lozoya et al. (2023)** | Australia | AI-generated synthetic mental health session data | Males associated with competence-related traits, females with warmth-related. Male-generated text: "He demonstrates strong analytical skills and a results-oriented approach." Female-generated text: "She shows great empathy and is always willing to lend an ear to others." |
| **Yang et al. (2024)** | USA | AI-generated medical reports | More severe disease predictions and higher costs for certain racial groups. For example: When presented with identical conditions, the model can diagnose HIV in Black patients, Tuberculosis in Asian patients, and cyst in White patients. |
| **Bozdag et al. (2024)** | Turkey | Legal language model outputs | Significant gender bias in word associations in legal contexts |
| **Xie et al. (2024)** | USA | AI-analyzed electronic health records | Worse seizure outcomes predicted for females, public insurance holders, and lower-income areas |

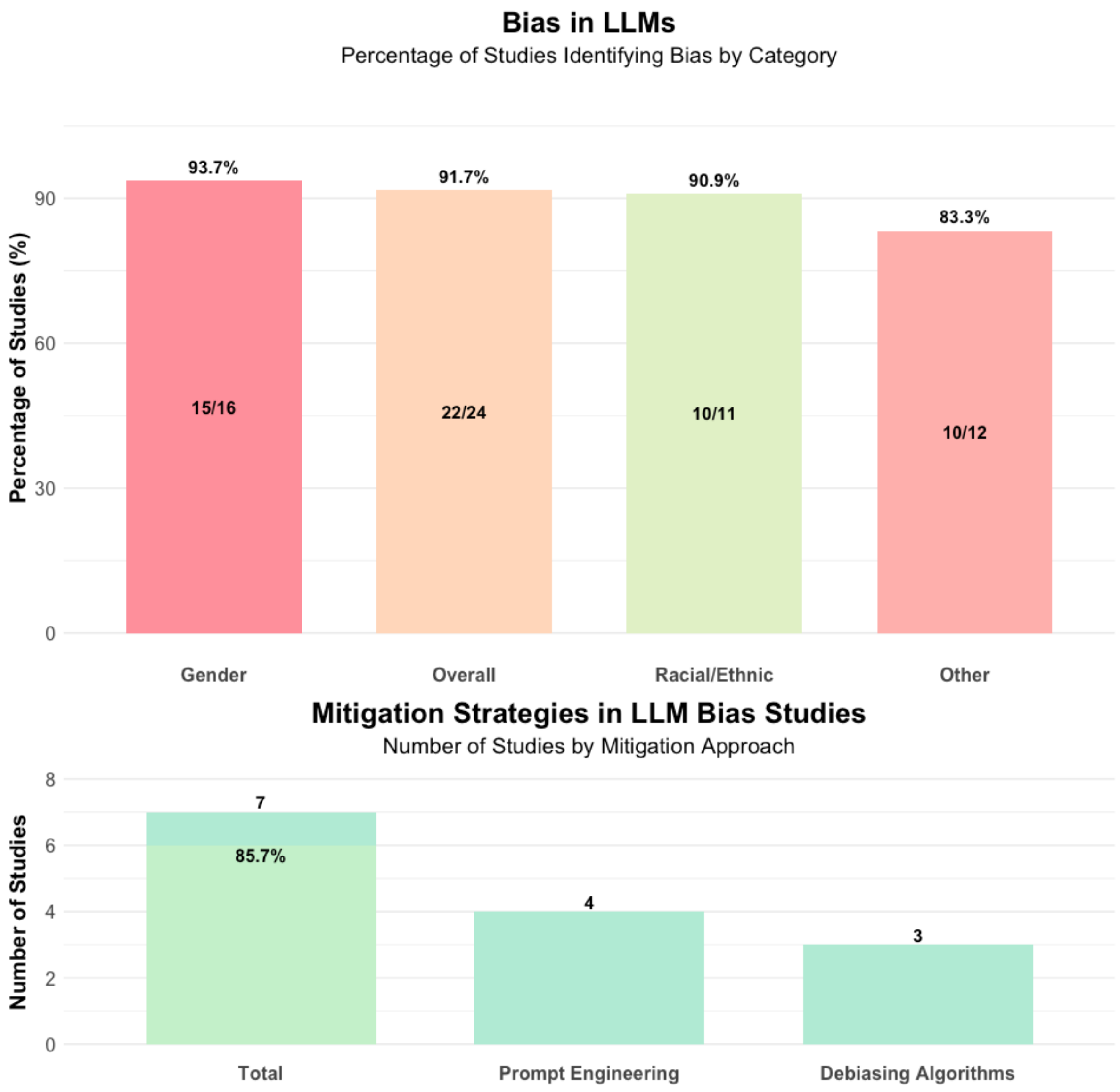

*The percentage in the lower "Total" bar represents the proportion of successful bias mitigation strategies (e.g., those reported as having successfully mitigated the bias)

**Figure S1**: A numeric overall analysis of the detected bias and mitigation strategies.
